# Oncologic, Functional, and Economic Outcomes of Transoral Robotic Surgery in HPV-Positive Oropharyngeal Squamous Cell Carcinoma: A Systematic Review and Meta-analysis of Pathology and Imaging-Guided Treatment Strategies

**DOI:** 10.64898/2026.01.11.26343884

**Authors:** Hamidreza Hajihosseni, Rosa Hajitaghizadeh, Hamid Reza Sharifi, Lin Dongpeng

## Abstract

**Background:** The incidence of human papillomavirus (HPV)-positive oropharyngeal squamous cell carcinoma (OPSCC) has increased substantially. Transoral robotic surgery (TORS) provides precise pathological staging that may facilitate treatment de-escalation; however, integrated evidence on oncological, functional, and economic outcomes remains limited.

**Objective:** To systematically evaluate oncological, functional, and economic outcomes of TORS-based treatment strategies in patients with HPV-positive OPSCC through systematic review and meta-analysis.

**Methods:** A systematic review and meta-analyses were conducted (PRISMA 2020). PubMed, Wiley, MDPI, Cureus, and MedRxiv were searched for studies published between 2019 and 2025. Meta-analyses were performed for 2-, 3-, and 5-year overall survival (OS), tracheostomy rates, long-term gastrostomy dependence, and postoperative bleeding using random-effects models.

**Results:** Twenty-two studies (20 clinical, 2 economic), comprising patients across 14 studies contributing to the quantitative meta-analysis. The pooled 2-, 3-, and 5-year overall survival (OS) rates were 86%, 96%, and 82%, respectively (with the 5-year OS reaching 92% in the consistent subgroup analysis). Functional preservation was excellent, with a remarkably low long-term gastrostomy dependence rate of 2% and a pooled tracheostomy rate of 9% (increasing to 11% after addressing study outliers). The pooled postoperative hemorrhage rate was 7%. Pathological upstaging following TORS frequently identified adverse features, enabling risk-adapted adjuvant therapy. De-escalation strategies were associated with substantial reductions in severe toxicity (Grade 3–5 toxicity dropping from 60% to 14% in key trials) while preserving quality of life. Economic modeling studies suggested that higher upfront surgical costs associated with TORS may be partially offset by reduced utilization of adjuvant therapy and lower rates of long-term treatment-related morbidity; however, these findings are based on limited available economic data.

**Conclusion:** TORS is an oncologically sound primary treatment for HPV-positive OPSCC, enabling accurate risk stratification and treatment de-escalation. Meta-analytic findings suggest favorable survival outcomes and low rates of major functional morbidity) 2% feeding tube dependence), supporting the potential value of TORS-based strategies in appropriately selected patient populations.

## 1. Introduction

Over the past two decades, the epidemiology of head and neck cancers has undergone a substantial shift. Oropharyngeal squamous cell carcinoma (OPSCC), historically associated with tobacco and alcohol exposure, has increasingly been driven by human papillomavirus (HPV) infection (1, 2). In the United States, this shift has reached epidemic proportions; between 1988 and 2004, the incidence of HPV-positive OPSCC rose by 225%, currently accounting for 80% to 90% of all new OPSCC diagnoses and surpassing cervical cancer as the most common HPV-related malignancy (3). Patients with HPV-positive OPSCC typically present at a younger age and demonstrate improved treatment response and survival outcomes compared with HPV-negative disease. As survival improves in this younger population, clinical priorities have expanded beyond disease control toward the preservation of long-term quality of life (QOL). Traditional treatment paradigms, historically reliant on definitive high-dose chemoradiotherapy (CRT), achieve effective oncologic control but are associated with substantial long-term toxicity, including dysphagia, xerostomia, and soft tissue fibrosis. This has fueled interest in treatment strategies aimed at reducing cumulative treatment-related morbidity, often framed within the concept of treatment de-escalation.

Transoral robotic surgery (TORS) has emerged as a surgical-first approach designed to balance oncologic efficacy with functional preservation. A principal advantage of TORS is the provision of definitive surgical pathology, allowing more accurate staging than imaging-based assessment alone (4, 5). The identification of adverse pathological features, such as positive margins and extranodal extension (ENE), enables refined risk stratification and supports tailored adjuvant therapy, including the potential for treatment de-escalation in appropriately selected patients (6–8). Despite the growing adoption of TORS, its optimal role within contemporary multidisciplinary treatment pathways remains incompletely defined. Much of the existing literature evaluates isolated aspects of TORS, focusing on survival outcomes or standalone economic modeling. In contrast, integrated assessments that concurrently examine oncologic control and the contribution of advanced intraoperative imaging to surgical adequacy (9) are relatively limited. Broader economic implications of TORS-based strategies (10, 11) are also sparsely addressed.

A comprehensive synthesis of these intersecting domains is necessary to clarify the clinical value of a TORS-first approach and to inform evidence-based de-escalation frameworks in HPV-positive OPSCC. Accordingly, this systematic review aims to provide an integrated evaluation of oncologic, functional, and economic outcomes associated with TORS-based treatment strategies in patients with HPV-positive OPSCC.

## 2. Methods and Materials

### 2.1 Inclusion Criteria

Eligibility criteria were defined a priori in accordance with the PICO framework **(Table 1)**. We included prospective and retrospective cohort studies as well as clinical trials evaluating transoral robotic surgery (TORS) in patients with head and neck squamous cell carcinoma (HNSCC), with specific emphasis on oropharyngeal squamous cell carcinoma (OPSCC). Studies were eligible if they enrolled adult patients (≥18 years) with histologically confirmed human papillomavirus (HPV)– positive OPSCC. Studies with mixed HPV status were also included. TORS was included as a surgical intervention both in primary and salvage treatment, intended as a curative procedure for HPV-positive OPSCC. Data from salvage TORS studies were primarily included for functional and morbidity outcomes and were distinguished from primary treatment results where applicable. Studies were required to report at least one of the following outcomes: local and/or regional disease control, surgical complications, treatment-related toxicities, or functional outcomes, including swallowing function, speech outcomes, and quality of life measures. Only articles published in English between 2019 and 2024 were included. This time restriction was applied to focus on studies reflecting contemporary robotic surgical platforms and to ensure alignment with the American Joint Committee on Cancer (AJCC) 8th edition staging system, which classifies HPV-positive OPSCC as a distinct clinical and biological entity.

**Table 1.** Eligibility criteria for study selection.

| Domain | Inclusion criteria | Exclusion criteria |
| --- | --- | --- |
| **Population** | Adult patients ($\geq 18$ years) | Paediatric populations |
| **Tumour site** | Oropharyngeal squamous cell carcinoma (OPSCC) | Non-oro-pharyngeal head and neck cancers (e.g., larynx, hypopharynx) |
| **Histology** | Squamous cell carcinoma | Non-squamous histology (e.g., salivary gland tumours) |
| **Biologic status** | Histologically confirmed HPV-positive disease (Mixed HPV status) | unreported HPV status |
| **Disease stage** | Resectable disease without distant metastasis | Unresectable disease or distant metastatic disease |
| **Intervention** | Transoral robotic surgery (TORS) in primary or salvage treatment | Open surgery, conventional non-robotic approaches, or non-surgical primary treatments |
| **Adjuvant therapy** | TORS alone or combined with adjuvant radiotherapy and/or chemoradiotherapy | Studies without a surgical component |
| **Treatment intent** | Curative intent (including upfront and salvage surgery) | Palliative treatment intent |
| **Outcomes reported** | Oncologic outcomes and/or surgical complications, treatment-related toxicities, functional outcomes (swallowing, speech, quality of life) | No extractable data on relevant clinical or functional outcomes |
| **Study design** | Prospective or retrospective cohort studies and clinical trials | Single case reports, animal studies, in vitro studies, systematic reviews |
| **Publication type** | Full-text, peer-reviewed articles | Conference abstracts without full text or non-peer-reviewed sources |
| **Language** | English | Non-English publications |
| **Publication period** | 2019–2024 | Published before 2019 |

### 2.2 Exclusion Criteria

Studies were excluded if they met any of the following criteria. Patients with non-oropharyngeal head and neck cancers (including laryngeal or hypopharyngeal malignancies), unresectable distant metastatic disease, or histological subtypes other than squamous cell carcinoma (e.g., salivary gland tumours) were excluded. Regarding study design, single case reports, animal studies, in vitro laboratory studies, and systematic reviews were excluded to avoid duplication of data. Studies in which transoral robotic surgery (TORS) was not employed as a surgical intervention, including those utilizing conventional open surgical approaches or exclusively non-surgical primary treatment modalities, such as radiotherapy or chemoradiotherapy, were excluded. Importantly, studies in which TORS was used for the management of recurrent or persistent disease were included, provided that relevant oncologic, clinical, or functional outcomes were reported. Additionally, studies were excluded if they failed to report patients’ HPV status or did not provide extractable data on relevant clinical, oncological, or functional outcomes. Conference abstracts without full-text publications and non–peer-reviewed or non-scientific sources were also excluded.

### 2.3 Search Strategy

PRISMA 2020 checklist and guidelines were applied in this article. A comprehensive literature search was conducted using internationally recognized electronic databases, including PubMed, Wiley Online Library, MDPI, Cureus, and medRxiv. The search strategy incorporated combinations of Medical Subject Headings (MeSH) terms and free-text keywords, including “oropharyngeal squamous cell carcinoma,” “human papillomavirus,” “transoral robotic surgery,” “TORS,” and “OPSCC.” These terms were selected to capture both broad head and neck oncology literature and studies specifically addressing HPV-positive OPSCC treated with robotic surgery, thus minimizing the risk of overlooking early-stage studies. The final literature search was conducted in November 2025. The complete electronic search strategy for all databases is provided in **Table 2**.

**Table 2.** search strategy for each database.

| Number of initial results | Role in search strategy | Database name |
| --- | --- | --- |
| 780 | Main reference for clinical articles and randomized trials | **PubMed** |
| 410 | Specialized source of articles on oropharyngeal surgery and anatomy | **Wiley Online Library** |
| 350 | Coverage of modern systematic reviews and practical implications | **MDPI** |
| 210 | Technical reports and new techniques in robotic surgery | **Cureus** |
| 168 | Identifying emerging data and scientific preprints | **medRxiv** |

### 2.4 Study Selection

A total of 1,918 records were identified through database searching. After removal of duplicates (n = 1,208), 710 records remained for title and abstract screening. Following this screening stage, 416 records were excluded due to irrelevance to the study topic. The full texts of 294 articles were assessed for eligibility. Of these, 184 articles were excluded based on predefined exclusion criteria (non-OPSCC population, absence of TORS intervention, insufficient outcome data, or non-peer-reviewed format). Ultimately, 20 clinical studies were included in the qualitative synthesis. In addition, two economic evaluation studies were included for cost analysis, resulting in a total of 22 studies included in the final review. The PRISMA 2020 flow diagram is presented in **Figure 1**.

**Figure 1.**
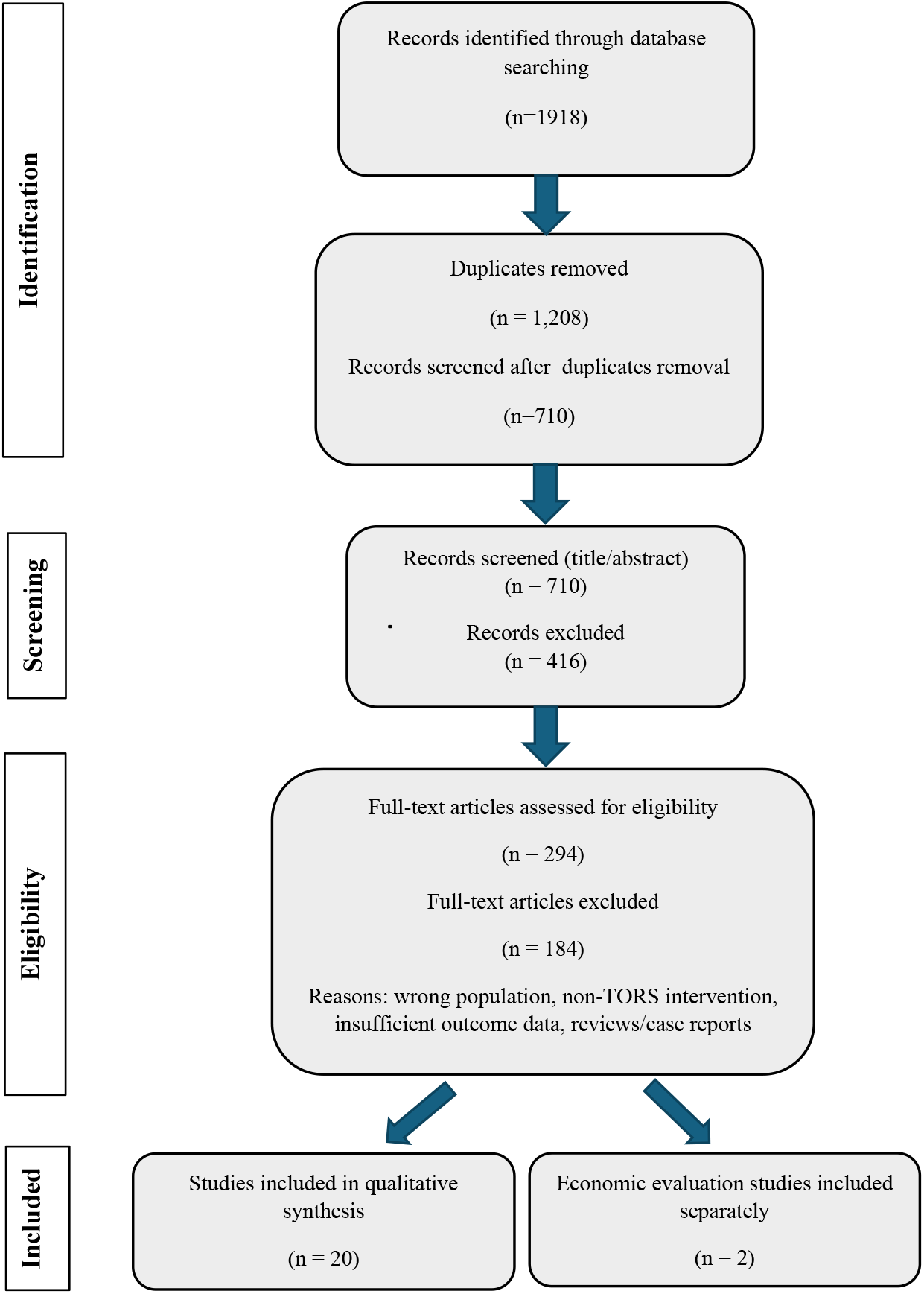
PRISMA 2020 flow diagram of study selection process

### 2.5 Quality Assessment of Included Studies

The methodological quality of the included studies was assessed using validated tools appropriate to each study design. Randomized trials and secondary analysis of randomized controlled trials were evaluated using the Cochrane Risk of Bias 2.0 (RoB 2.0) tool. Non-randomized interventional studies were assessed using the ROBINS-I and MINORS instruments where appropriate. Observational cohort studies were evaluated using the Newcastle–Ottawa Scale (NOS), while diagnostic accuracy studies were appraised using the QUADAS-2 tool. Case series were assessed using the Joanna Briggs Institute (JBI) Critical Appraisal Checklist. Overall, most included studies demonstrated high methodological quality. Most cohort studies achieved NOS scores of 8–9 stars, indicating low risk of bias. Randomized and secondary trial analysis were judged to have low risk of bias using RoB 2.0. Non-randomized interventional studies showed moderate to high methodological quality, while diagnostic studies demonstrated acceptable risk profiles. Case series consistently met all JBI quality criteria. A summary of the quality assessment is presented in **Table 3**.

**Table 3.**
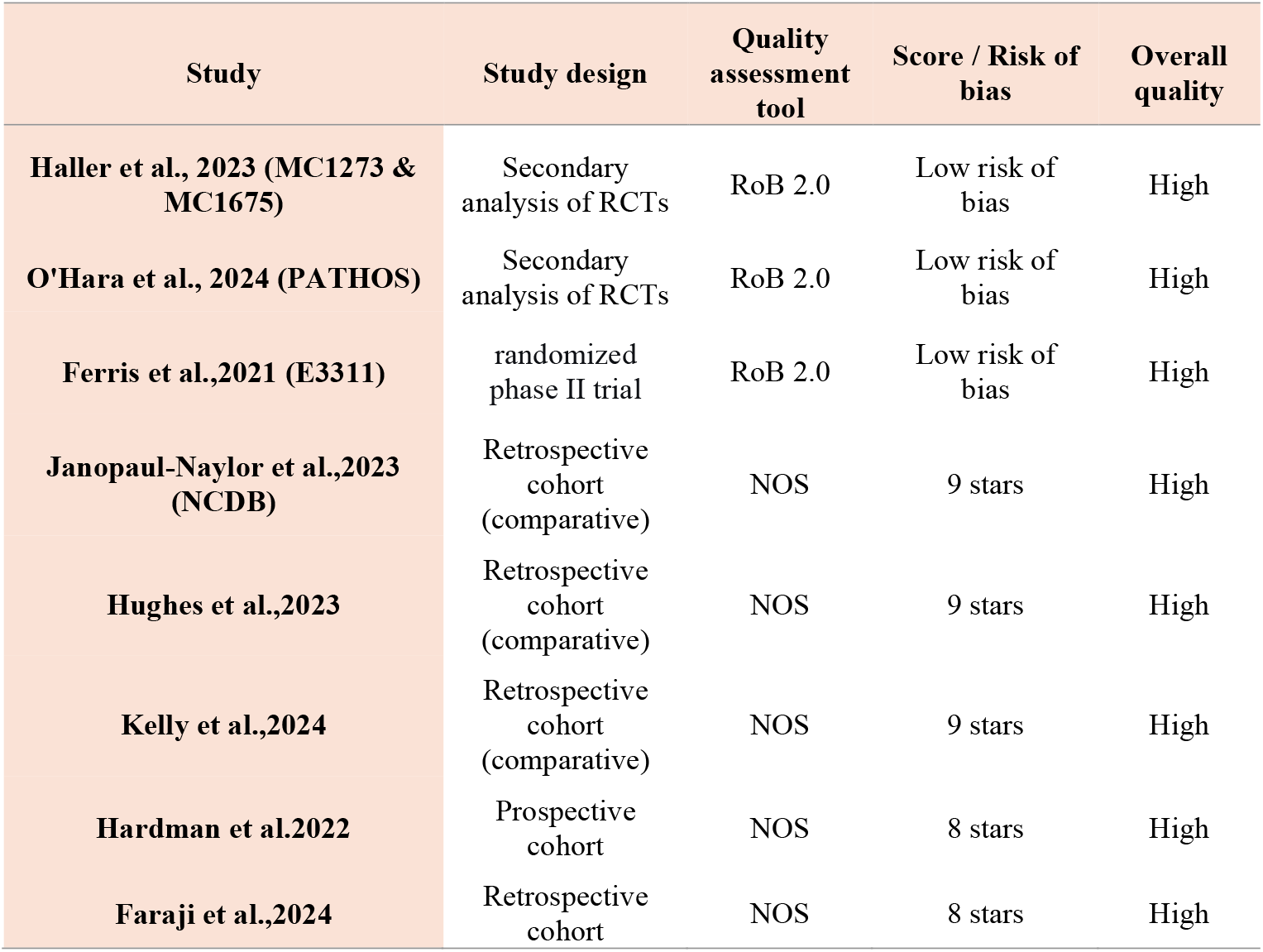
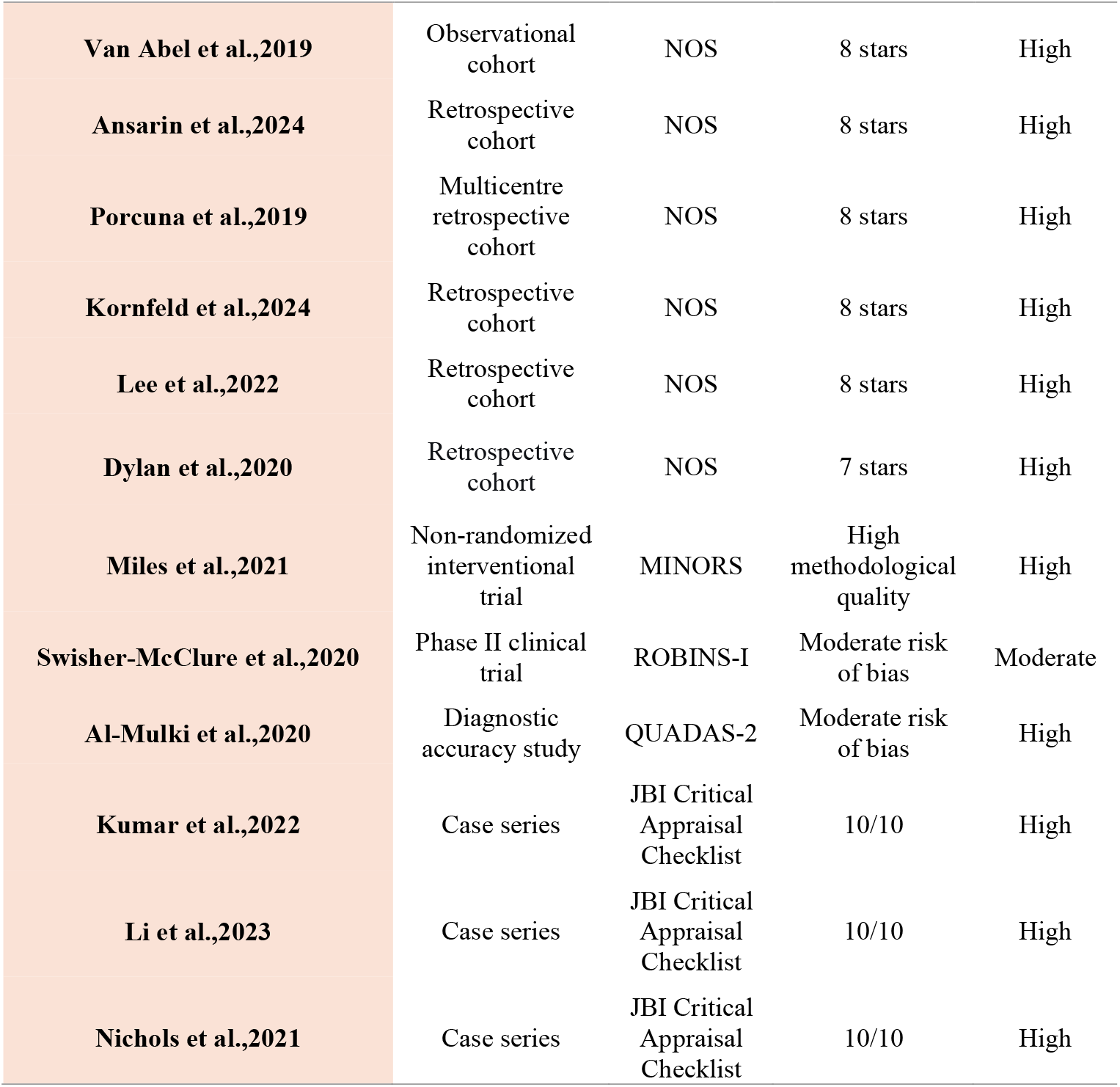
Quality assessment.

Despite the generally high methodological quality of included studies, most evidence was derived from observational cohorts, which are inherently prone to selection bias and residual confounding. Heterogeneity in study design, outcome definitions, and follow-up limited quantitative synthesis for some endpoints. Non-randomized interventional studies demonstrated moderate risk of bias, and variability in HPV assessment and staging systems, which may have affected comparability. Economic analysis were limited and may not fully reflect institutional cost differences.

### 2.6 Data Extraction

At this stage, data were extracted from 22 eligible studies using a standardized data extraction form developed in Microsoft Excel. To ensure accuracy and reliability, data extraction was conducted independently by two reviewers, with discrepancies resolved through discussion and consensus. Digital reference management software (EndNote) was used to organize citations, and Good Notes was employed for full-text annotation and note-taking.

Extracted data were systematically categorized into five predefined domains. Study characteristics and patient demographics included first author, year of publication, study design, sample size, mean or median age, sex distribution, and smoking history, quantified in pack-years as a potential confounding variable. Diagnostic and surgical variables comprised human papillomavirus (HPV) status and method of confirmation (p16 immunohistochemistry, polymerase chain reaction, or in situ hybridization), transoral robotic surgery (TORS) platform, primary tumour site, and the timing and extent of neck dissection. Pathologic features included surgical margin status, tumour staging according to the American Joint Committee on Cancer (AJCC) 7th or 8th edition, and the presence of high-risk pathological features such as lymphovascular invasion and extranodal extension. Oncologic outcomes and adjuvant treatments encompassed overall survival, disease-specific survival, progression-free survival, recurrence-free survival, patterns of recurrence, and the use of adjuvant radiotherapy or chemoradiotherapy, including treatment de-escalation strategies in low-risk patient cohorts. Functional and quality-of-life outcomes included scores from the MD Anderson Dysphagia Inventory (MDADI) and the Functional Oral Intake Scale (FOIS), long-term feeding tube dependence, and postoperative complications such as hemorrhage and the need for tracheostomy. Economic outcomes, when reported, included direct and indirect costs to inform cost-effectiveness analysis.

### 2.7 Data Synthesis and Statistical Analysis

Data synthesis followed the PRISMA 2020 guidelines using a combined quantitative and narrative approach. All statistical meta-analyses were conducted using Review Manager (RevMan) [Computer program], Version 5.4 (The Cochrane Collaboration, 2020). Studies were initially grouped by treatment strategy, including TORS alone, TORS with adjuvant therapy, and comparative studies, to facilitate descriptive synthesis. Quantitative meta-analysis was performed for outcomes with sufficient data, including overall survival (OS) at 2-, 3-, and 5-year intervals, postoperative tracheostomy rates, long-term feeding tube dependence, and postoperative hemorrhage. Pooled estimates and their 95% confidence intervals (CIs) were calculated using random-effects models (DerSimonian and Laird) to account for anticipated clinical and methodological heterogeneity. Between-study heterogeneity was assessed using the Cochran Q test and quantified by the I^2^ statistic, where I^2^ > 50% indicated substantial heterogeneity. To investigate the sources of high heterogeneity and ensure the robustness of the results, sensitivity analysis were conducted using an outlier-removal approach. Studies were categorized into ‘Consistent’ and ‘Outlier’ subgroups to identify studies that disproportionately contributed to statistical heterogeneity. Additionally, key clinical characteristics, including smoking history and advanced tumor stage (T3–T4), were extracted and summarized in the Characteristics of Included Studies table (Table 5) to provide a comprehensive clinical context for the included populations. For outcomes not suitable for quantitative pooling—such as quality-of-life (QoL) measures, MD Anderson Dysphagia Inventory (MDADI) scores, and economic outcomes—a narrative synthesis was provided. Formal assessment of publication bias via funnel plots was not performed due to the limited number of studies (*n* < 10) per outcome, in accordance with Cochrane recommendations.

## 3. Result

### 3.1 Study Characteristics and Data Synthesis

A total of 22 studies met the inclusion criteria, consisting of 20 clinical reports (1–9, 12–22) and two economic models (10, 11). The synthesis incorporates a total of 7,042 patients, stratified into two primary data streams for analytical clarity. First, large-scale registry data were extracted from the National Cancer Database (NCDB), encompassing 3,639 patients treated at Commission on Cancer-accredited facilities (5). Second, a collective cohort of 3,403 patients was synthesized from the remaining 19 clinical studies, comprising Phase II and III prospective trials (e.g., E3311, AVOID, MC1273, and MC1675) (7, 8, 19), the international RECUT cohort (18), and various institutional series (1–4, 9, 13–17, 20, 22). By separating registry-based data from trial-specific and institutional cohorts, this synthesis offers complementary perspectives: it highlights broad real-world trends while also providing high-granularity clinical outcomes following transoral surgery **(Table 4)**. Although the total synthesized population comprised 7,042 patients, large registry-based datasets (e.g., NCDB, n = 3,639) were analyzed independently and not combined with smaller clinical cohorts in a weighted analysis. This approach was adopted to preserve the interpretability of trial-level and institutional outcomes and to prevent dominance of large-scale observational data. The main characteristics of the included clinical studies, including study design, patient demographics, tumour stage, and adjuvant treatment patterns, are summarized in **Table 5**.

**Table 4.** Study Characteristics and Data Sources Included in the Synthesis.

| Data stream | Study type | Included studies | Number of studies (n) | Patients (n) | Data source description |
| --- | --- | --- | --- | --- | --- |
| **Registry-based data** | Registry analysis | NCDB | 1 | 3,639 | National Cancer Database; Commission on Cancer–accredited centres |
| **Prospective clinical trials** | Phase II–III trials | E3311, AVOID, MC1273, MC1675 | 4 | Patients included in pooled cohort | Protocol-driven prospective studies |
| **International cohort** | Multicentre cohort | RECUT | 1 | Patients included in pooled cohort | International multicentre dataset |
| **Institutional series** | Retrospective cohorts | Institutional reports | 14 | Patients included in pooled cohort | Single- and multi-institutional series |
| **Clinical studies (total)** | Mixed designs | Clinical reports | 20 | 3,403 | Combined trial-based and institutional cohorts |
| **Economic models** | Cost-effectiveness analysis | Economic studies | 2 | — | Decision-analytic and cost models |
| **Overall synthesis** | — | All included studies | 22 | **7,042** | Combined registry, clinical, and economic data |

**Table 5.** Characteristics of Included Clinical Studies.

| **Author (Year)** | **Study Design** | **Total N** | **HPV + (%)** | **Age Mean/Med** | **Male (%)** | **Smoking History** | **Neck Dissection (%)** | **Advanced T stage (T3–T4) (%)** | **Adjuvant RT\* / CRT\* (%)** | **Follow-up (Months)** |
| --- | --- | --- | --- | --- | --- | --- | --- | --- | --- | --- |
| **Miles et al. (2021)** | Phase II Trial | 54 | 100 % | 59.0 | 100.0 | <20 pack-years (5.5%) | 100.0 | 0.0 | 27.7% / 25.9% | 43.9 |
| **Faraji et al. (2024)** | Retrospective | 161 | 100 % | 64.0 | 98.0 | ≥10 pack-years (57.0%) | NR* | 13.0 | 23.6% / 47.8% | 67.2 |
| **Haller et al. (2023)** | Phase II/III | 219 | 100 % | 58.9 | 89.0 | Any history (33.0%) | 100.0 | 13.7 | NR | 1.0 (acute)* |
| **James T et al. (2024)** | RCT Cohort | 508 | 100 % | 58.3 | 76.8 | Ex/Current (43.8%) | 98.4 | 3.5 | NR | 60.0 |
| **Lee et al. (2022)** | Retrospective | 37 | 100 % | 59.0 | 94.6 | Never-smoker (70.3%) | 100.0 | 27.0 | NR | 45.6 |
| **Li et al. (2023)** | Retrospective | 83 | 30.1 %* | 57.0 | 85.5 | History (54.2%) | 83.1 | 0.0 | 6.0% / 21.7% | 29.5 |
| **Roden et al. (2020)** | Retrospective | 286 | 100 % | 60.0 | 87.0 | History (57.4%) | 100.0 | 7.8 | 84.5% (Total RT) | 38.8 |
| **Nichols et al. (2021)** | Retrospective | 48 | 100 % | 61.2 | 83.0 | Ex/Current (79.0%) | NR | 0.0 | 79.0% (Indicated) | 26.4 |
| **Hardman et al. (2022)** | Int. Cohort | 278 | NR | 61.0 | 79.9 | Ex/Current (73.6%) | 100.0 | NR | NR | 38.5 |

For the quantitative synthesis (meta-analysis), a subset of 14 clinical studies provided sufficient granular data for pooling. These studies were meta-analyzed across six key outcomes, including oncologic survival (2, 3, and 5-year OS) and surgical morbidity (post-operative bleeding, tracheostomy dependency, and long-term feeding tube rates). The pooled results are presented in the following sections and illustrated in **Figures 2 through 7**.

**Figure 2.**
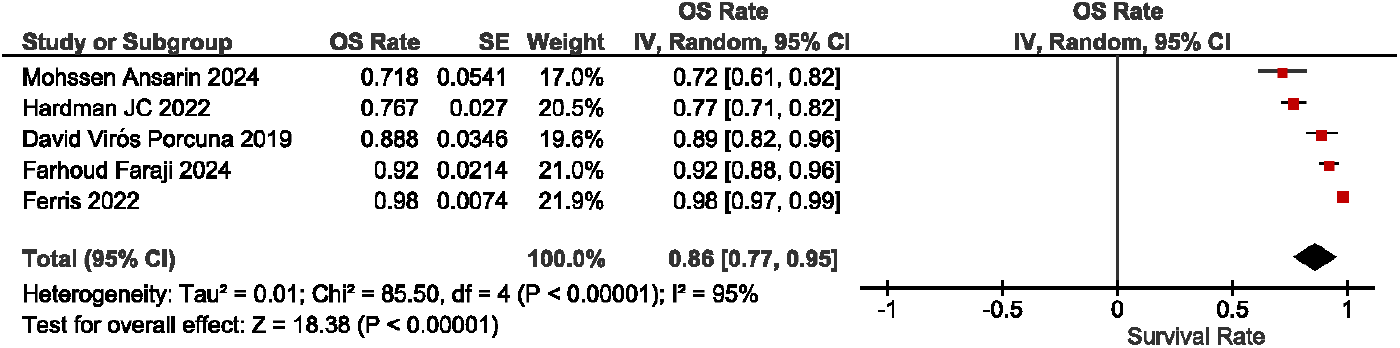
Forest plot of comparison: 2-year overall survival

### 3.2 Oncologic Outcomes

The quantitative synthesis of oncologic outcomes revealed robust survival rates for the 2-year follow-up interval. The pooled 2-year overall survival (OS) rate was 0.86 (95% CI: 0.77–0.95), with high inter-study heterogeneity (I^2^ = 95%, P < 0.00001; **Figure 2**). While individual study data reported 2-year OS between 72% (2) and 98% (6), prospective benchmarks further solidified these results, with high-performing cohorts achieving up to 98% survival (19).

Mid-term oncologic control remained excellent, as the pooled 3-year OS rate was estimated at 0.96 (95% CI: 0.92–0.99) with moderate-to-high heterogeneity (I^2^ = 84%; **Figure 3**). In addition to overall survival, individual studies reported a 3-year Recurrence-Free Survival (RFS) of 90.7% and a locoregional control rate of 97.5% (3). Interestingly, although 57.4% of the synthesized population had a smoking history (mean 14.8 pack-years), meta-regression and subgroup data from individual trials showed no significant difference in 3-year RFS between never-smokers and smokers (92% vs. 89.8%; P = 0.85) (3).

**Figure 3.**
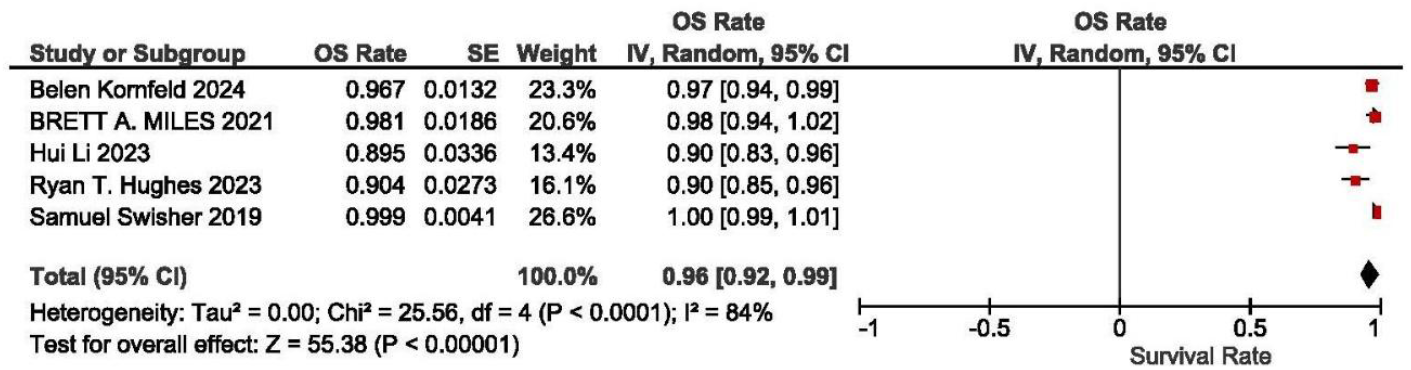
Forest plot of comparison: 3-year overall survival

For long-term outcomes, the total pooled 5-year OS rate was 0.82 (95% CI: 0.70–0.94). To address the significant heterogeneity (I^2^ = 98%), a subgroup analysis was performed. After excluding outlier data, the 5-year OS rate in the “Consistent Studies” subgroup was 0.92 (95% CI: 0.90–0.95) with a remarkably high level of homogeneity (I^2^ = 0%; **Figure 4**). These results are summarized in Table 6, covering overall survival, recurrence-free survival, and disease-specific outcomes.

**Table 6.** Summary of Oncologic Outcomes Following Transoral Surgery.

| Study / Data source | Study design | Population characteristics | Oncologic outcome | Value |
| --- | --- | --- | --- | --- |
| **Hughes et al.** | Retrospective cohorts | Mixed-risk OPSCC | 2-year OS | 90.4% |
| **Miles et al.** | Retrospective cohorts | Mixed-risk OPSCC | 2-year OS | 98.1% |
| **Ferris et al., E3311** | Prospective Phase II | High-risk group | 2-year OS | 96.3% |
|  |  | Low-risk group |  | 100% |
| **Roden et al.** | Retrospective | HPV+ OPSCC | 3-year RFS | 90.7% |
|  |  |  | Locoregional control (3-year) | 97.5% |
|  |  | Never-smokers | 3-year RFS | 92.0% |
|  |  | Ever-smokers (mean 14.8 PY) | 3-year RFS | 89.8% |
|  |  | Smoking comparison | P-value | 0.85 |

**Figure 4.** Forest plot of comparison: 5-year overall survival

### 3.3 Pathological Findings and Surgical Margins

Analysis of pathological findings across the included studies demonstrated significant shifts between clinical and pathological staging. Surgical pathology led to the upstaging of clinical T1–T2 disease to pT3–T4 in multiple cohorts (4, 19). Beyond standard staging, advanced pathological assessment revealed lymphovascular invasion (LVI) in 38% and perineural invasion (PNI) in 19% of cases (3).

A notable discrepancy was observed regarding surgical margins: expert-led prospective trials reported a positive surgical margin (PSM) rate of only 3.3% (19), whereas the broader registry-based NCDB analysis showed a significantly higher rate of 14.7% (5). Furthermore, active smoking was significantly associated with an increased risk of extranodal extension (ENE) and PSM (3). In the prospective setting, 34% of ENE cases were classified as “subtle” (≤ 1 mm), which facilitated the de-escalation of adjuvant therapy in these patients (19). These pathological characteristics are summarized in **Table 7**.

**Table 7.**
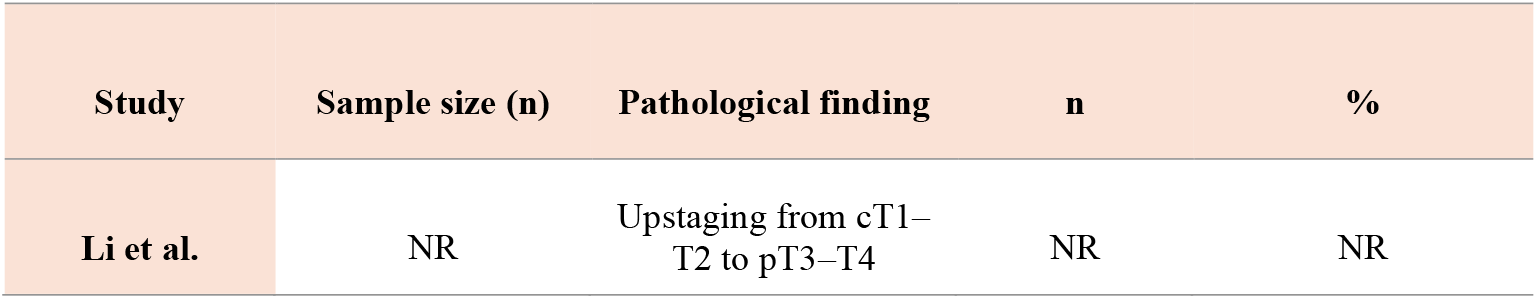
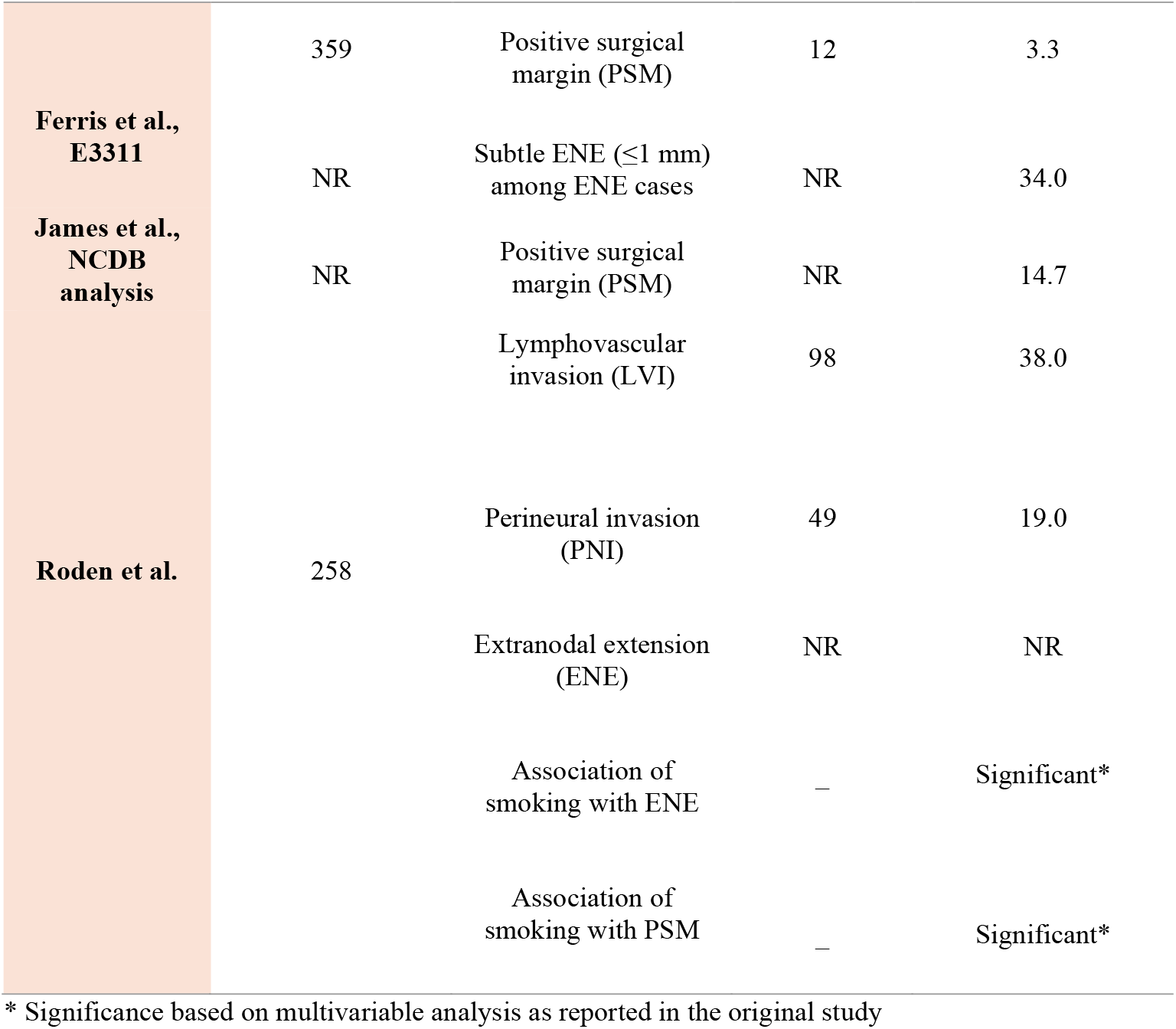
Pathological Findings and Surgical Margin Status Across Included Studies.

### 3.4 Functional Outcomes and Morbidity

The quantitative synthesis of surgical complications and functional recovery highlighted the safety profile of transoral approaches. The pooled postoperative hemorrhage rate was 0.07 (95% CI: 0.05– 0.10). Although individual reports showed a wide range between 0.8% and 14.9% (7, 13, 19), subgroup analysis of consistent studies confirmed a robust bleeding rate of 7% with no heterogeneity (I^2^ = 0%; **Figure 5**).

**Figure 5.**
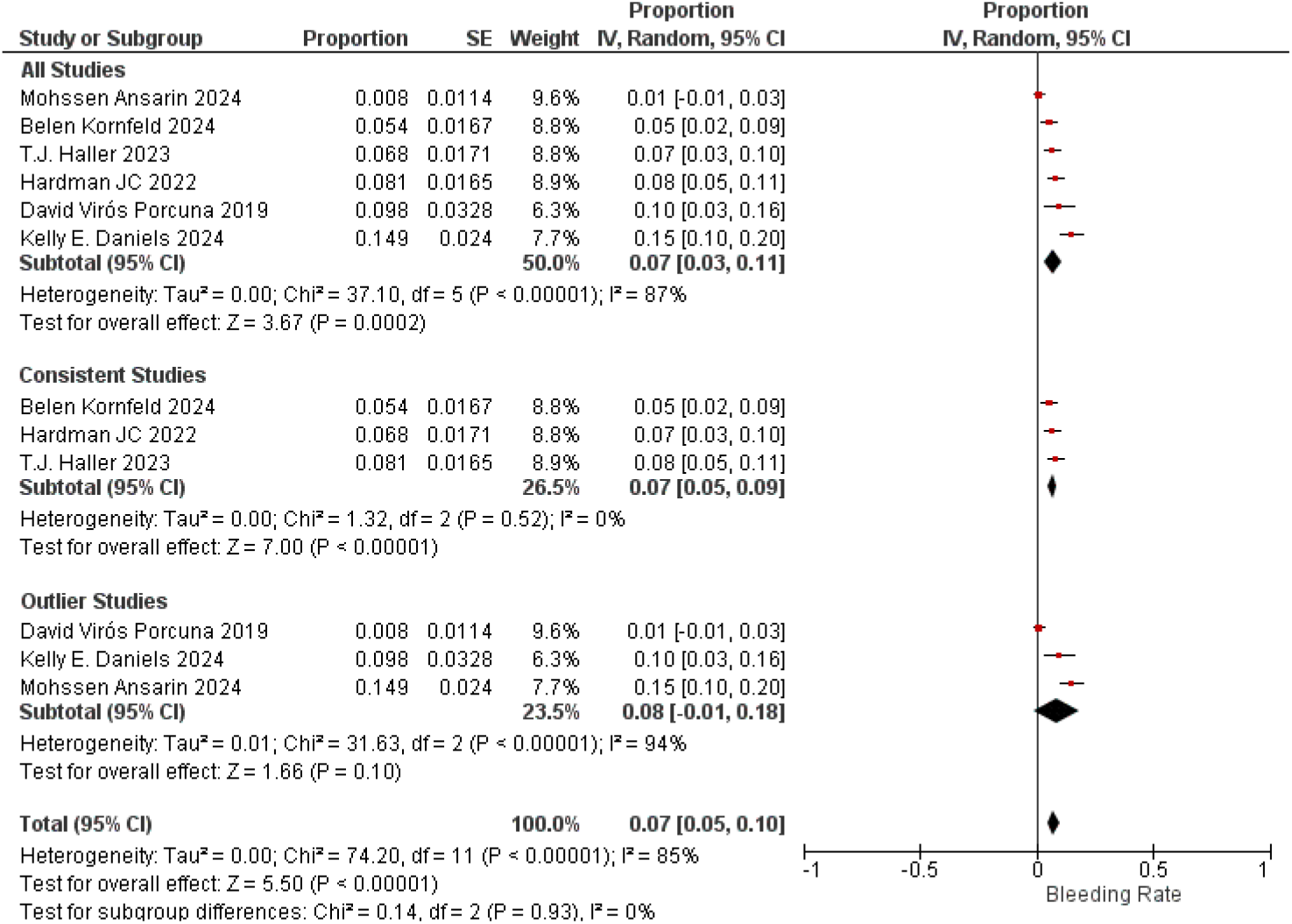
Forest plot of comparison: Postoperative bleeding

Airway and nutritional independence were generally well preserved across the included studies. The pooled tracheostomy rate was 0.09 (95% CI: 0.05–0.12), with a consistent subgroup rate of 11% after addressing study outliers (**Figure 6**).

**Figure 6.**
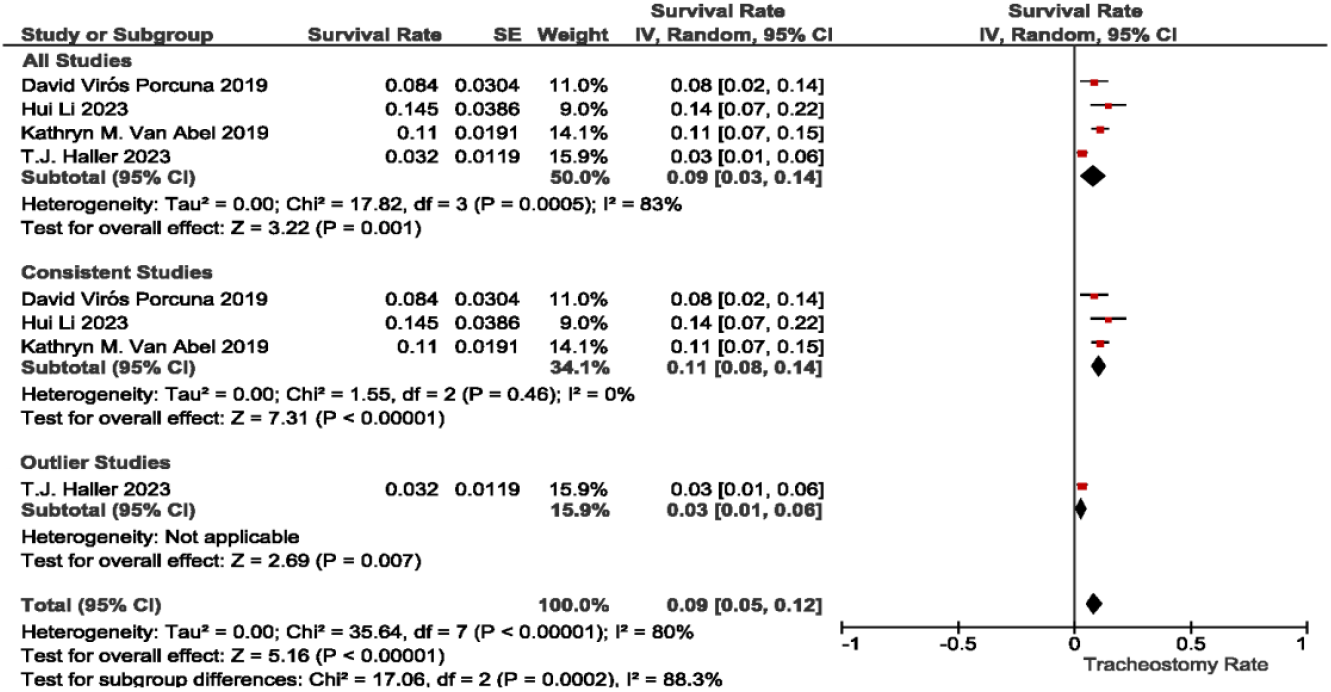
Forest plot of comparison: Tracheostomy Rate

Regarding long-term nutritional support, the meta-analysis revealed a remarkably low gastrostomy dependence rate of only 0.02 (95% CI: 0.00–0.03) with absolute homogeneity (I^2^ = 0%; **Figure 7**). This confirms that while some studies reported dependence below 5% (8, 19, 21), the aggregate evidence suggests an even more favorable outcome.

**Figure 7.**
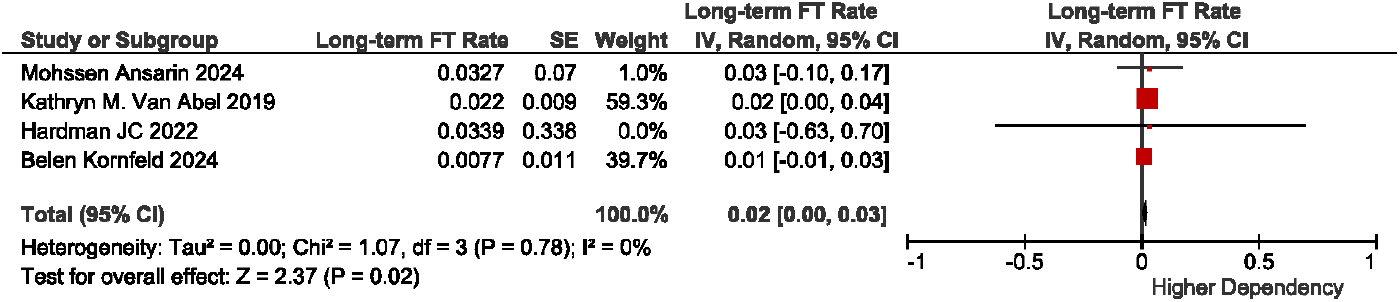
Forest plot of comparison: Feeding tube

Furthermore, de-escalated adjuvant protocols significantly reduced treatment-related toxicity. Severe Grade 3–5 toxicity dropped to 14% in the 50 Gy arm, compared to 60% in the standard high-risk arm (19). Quality of life and swallowing scores in these de-escalated groups typically returned to baseline levels within six months, reinforcing the functional benefits of this treatment paradigm. These outcomes are summarized in **Table 8**.

**Table 8.** Functional Outcomes and Treatment-Related Morbidity After Transoral Surgery.

| Study | Study design | Outcome domain | Outcome measure | Value |
| --- | --- | --- | --- | --- |
| **Haller et al.** | Retrospective (pooled prospective trials) | Surgical morbidity | Postoperative hemorrhage | 0.5–15.1% |
| **Kornfeld et al.** | Retrospective single-centre | Surgical morbidity | Postoperative hemorrhage | Included in range |
| **Swisher-McClure et al., AVOID** | Prospective Phase II | Quality of life | Swallowing/QoL recovery | Returned to baseline $\leq 6$ months |
| **Ferris et al., E3311** | Prospective Phase II RCT | Treatment-related toxicity | Grade 3–5 toxicity (50 Gy arm) vs Grade 3–5 toxicity (standard high-risk arm) | 14% vs 60% |
| | | Quality of life | Swallowing/QoL recovery | Returned to baseline ≤6 months |
| **Viros Porcuna et al.** | Multicentre retrospective | Functional outcome | Long-term gastrostomy dependence | <5% |

### 3.5 Economic Evaluation

Direct surgical costs for TORS remained slightly higher than those for TLM (10) **(Table 9)**. However, the economic viability of the robotic approach is supported by a lower probability of requiring expensive adjuvant chemoradiotherapy (52.7% vs. 62.9%) (10). Modelling indicates that TORS is cost-saving when adjuvant therapy requirements remain below 59.7% or when margin revision rates for alternative techniques exceed 29% (10, 11) **(Table 10)**.

**Table 9.** direct costs of surgery.

| Surgical Technique | Mean Cost (USD) |
| --- | --- |
| **Non robotic surgery (TLM)** | \$58,334.93 |
| **Robotic surgery (TORS)** | \$61,998.25 |
| **Cost difference** | +\$3,663.33 (TORS higher) |

**Table 10.**
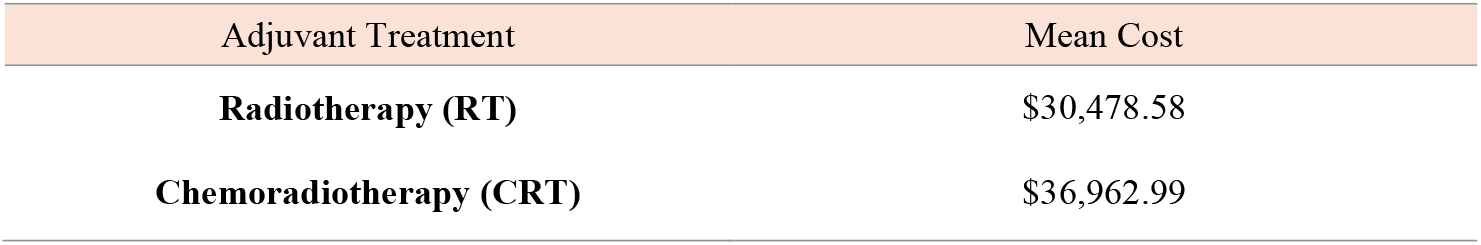
Costs of adjuvant therapy.

## 4. Discussion

This systematic review and meta-analysis suggest that Transoral Robotic Surgery (TORS) may represent an effective primary treatment approach in selected patients. Our findings underscore excellent survival outcomes; specifically, the meta-analysis showed a pooled 2-year OS of 86% (95% CI: 0.77–0.95). Notably, in the subset of studies reporting longer follow-ups, the 3-year and 5-year OS rates remained excellent at 96% (**Figure 3**) and 92% in consistent studies (**Figure 4**), respectively. These findings are consistent with previous reports in which survival rates exceeded 95% and 3-year progression-free survival (PFS) ranged from 88% to 96% (2, 3, 19). The inclusion of the prospective ECOG-ACRIN E3311 trial serves as a pivotal validation, shifting the focus from surgical feasibility to intelligent treatment de-escalation. By providing definitive surgical pathology, TORS enables the omission of adjuvant therapy in low-risk cases or a substantial reduction in radiation dose to 50 Gy for intermediate-risk patients, achieving a 2-year PFS of 94.9% (19).

This granular pathological assessment highlights a key limitation of imaging-based staging and reinforces the role of upfront surgery in tailoring postoperative management. A critical aspect of this review was the evaluation of smoking status in relation to surgical outcomes. While smoking is a well-established negative prognostic factor in radiotherapy-based treatments, no significant difference in recurrence-free survival (RFS) was observed between smokers and non-smokers treated with TORS (3). This “hypoxia bypass” effect suggests that the physical removal of the tumor mass mitigates the oxygen-dependent resistance mechanisms inherent to non-surgical modalities. However, active smoking remains biologically relevant. As nicotine acts as an immunosuppressant by inhibiting T-cell and natural killer (NK) cell responses, this may explain the higher incidence of extranodal extension (ENE) and positive surgical margins observed in smoking populations (3). The value of surgical pathology is further demonstrated by its ability to uncover occult adverse features such as ENE, reported in up to 41% of node-positive cases (14). Importantly, the identification of “subtle” ENE ($\le 1$ mm) suggests that these patients may not require the full toxic burden of chemoradiotherapy (19). Nonetheless, clinicians must remain vigilant regarding the “triple-modality trap,” defined as the combined use of surgery, radiation, and chemotherapy. To avoid this scenario, careful patient selection is essential; patients with unfavorable anatomy should be triaged toward definitive chemoradiotherapy to prevent compounded morbidity (1, 6, 16).

Functional preservation remains a cornerstone of the TORS paradigm. Our meta-analysis revealed a pooled postoperative hemorrhage rate of 7% (**Figure 5**), providing a robust aggregate for the 0.8% to 14.9% range observed in individual trials (7, 13, 19). Airway and nutritional independence were also confirmed, with a pooled tracheostomy rate of 9–11% (**Figure 6**) and a remarkably low long-term gastrostomy dependence rate of only 2% (**Figure 7**), further strengthening individual study reports of dependence below 5% (8, 19, 21). Improved surgical proficiency and de-escalated treatment protocols may contribute to improved economic efficiency, although available cost data remain limited. As reductions in Grade 3–5 adverse events from 60% to 14% (19) minimize costly downstream consequences such as hospital readmissions. By maximizing quality-adjusted life years and facilitating a faster return to baseline function, TORS may represent a potentially high-value intervention within the evolving framework of treatment de-escalation and toxicity reduction strategies.

## 5. Conclusion

TORS appears to be an oncologically effective primary treatment option for selected patients with HPV-positive oropharyngeal squamous cell carcinoma, with favorable survival outcomes reported across included studies.The clinical utility of this approach lies in its ability to provide definitive surgical pathology, enabling a transition from imaging-based assumptions to precise risk stratification. By identifying granular pathological features such as subtle extranodal extension (ENE), lymphovascular invasion (LVI), and perineural invasion (PNI), TORS facilitates the implementation of intelligent treatment de-escalation strategies.

Crucially, across the included studies, smoking status was not associated with inferior survival outcomes in patients treated with TORS, likely because surgical resection bypasses hypoxia-related resistance mechanisms inherent to non-surgical therapies. Although initial surgical costs are higher, this approach demonstrates substantial potential for cost-effectiveness by reducing severe morbidity and decreasing the institutional burden associated with adjuvant chemoradiotherapy.although current evidence remains limited and requires validation through larger prospective economic studies. However, these benefits are contingent upon careful patient selection to avoid the triple-modality trap and the maintenance of a high level of surgical proficiency.

## Data Availability

All data analyzed during this study are included in this published article (and its supplementary information files). The data were extracted from previously published studies available in the public domain.

## 6. Declarations

The authors declare that they have no known competing financial interests or personal relationships that could have appeared to influence the work reported in this paper.

## 7. Author Contributions

Conceptualization, H.H.; PICO formulation and protocol development, R.H.; search strategy design, H.H. and R.H.; literature search and database retrieval, H.S.; study screening and selection, H.H. and R.H.; data extraction and curation, H.S.; quality assessment and risk of bias evaluation, H.H.; data synthesis and formal analysis (meta-analysis), H.H.; statistical validation and meta-analysis review, L.D.; writing—original draft preparation, H.H.; writing—review and editing, H.H. and L.D.; supervision, L.D.; economic outcomes analysis, H.S.

All authors have read and agreed to the published version of the manuscript.

## References

1. Li H, Zhang X, Chen W, Zhang Q, Li Q, Chen S, et al. Analysis of T1‐T2 stage oropharyngeal squamous cell carcinoma treated with transoral robotic surgery. Laryngoscope Investigative Otolaryngology. 2023;8(1):103–12.

2. Hughes RT, Levine BJ, May N, Shenker RF, Yang JH, Lanier CM, et al. Survival and swallowing function after primary radiotherapy versus transoral robotic surgery for human papillomavirus-associated oropharyngeal squamous cell carcinoma. ORL. 2023;85(5):284–93.

3. Roden DF, Hobelmann K, Vimawala S, Richa T, Fundakowski CE, Goldman R, et al. Evaluating the impact of smoking on disease‐specific survival outcomes in patients with human papillomavirus– associated oropharyngeal cancer treated with transoral robotic surgery. Cancer. 2020;126(9):1873–87.

4. Ansarin M, Pietrobon G, Tagliabue M, Mossinelli C, Ruju F, Maffini F, et al. Salvage transoral robotic surgery in recurrent oropharyngeal carcinoma: a single-centre retrospective study. European Archives of Oto-Rhino-Laryngology. 2024;281(6):3167–77.

5. Janopaul‐Naylor JR, Rupji M, Tobillo RA, Lorenz JW, Switchenko JM, Tian S, et al. Ninety‐day mortality following transoral robotic surgery or radiation at Commission on Cancer‐accredited facilities. Head & neck. 2023;45(3):658–63.

6. Miles BA, Posner MR, Gupta V, Teng MS, Bakst RL, Yao M, et al. De-escalated adjuvant therapy after transoral robotic surgery for human papillomavirus-related oropharyngeal carcinoma: The Sinai robotic surgery (SIRS) trial. The oncologist. 2021;26(6):504–13.

7. Haller T, Yin X, O’Byrne T, Moore E, Ma D, Price K, et al. 30-day morbidity and mortality after transoral robotic surgery for human papillomavirus (HPV) associated oropharyngeal squamous cell carcinoma: A retrospective analysis of two prospective adjuvant de-escalation trials (MC1273 & MC1675). Oral Oncology. 2023; 137:106248.

8. Swisher-McClure S, Lukens JN, Aggarwal C, Ahn P, Basu D, Bauml JM, et al. A phase 2 trial of alternative volumes of oropharyngeal irradiation for de-intensification (AVOID): omission of the resected primary tumour bed after transoral robotic surgery for human papilloma virus–related squamous cell carcinoma of the oropharynx. International Journal of Radiation Oncology* Biology* Physics. 2020;106(4):725–32.

9. Al-Mulki K, Hamilton J, Kaka AS, Boyce BJ, Baddour HM, El-Deiry M, et al. Narrowband imaging for p16+ unknown primary squamous cell carcinoma prior to transoral robotic surgery. Otolaryngology–Head and Neck Surgery. 2020;163(6):1198–201.

10. Parimbelli E, Soldati F, Duchoud L, Armas GL, de Almeida J, Broglie M, et al. Cost-utility of two minimally invasive surgical techniques for operable oropharyngeal cancer: transoral robotic surgery versus transoral laser microsurgery. BMC health services research. 2021;21(1):1173.

11. Tang Y, Dou B. Cost-effectiveness analysis of robotic surgery in healthcare for older individuals: a systematic review based on randomized controlled trials. Frontiers in Public Health. 2025; 13:1614654.

12. O’Hara JT, Hurt CN, Ingarfield K, Patterson JM, Hutcheson K, Canham JE, et al. Transoral laser or robotic surgery outcomes for oropharyngeal carcinoma: secondary analysis of the PATHOS randomized clinical trial. JAMA Otolaryngology–Head & Neck Surgery. 2024;150(11):1002–11.

13. Kornfeld B, Taha A, Kyang L, Sim H-w, Dewhurst S, McCloy R, et al. Oncological outcomes post transoral robotic surgery (TORS) for HPV-associated oropharyngeal squamous cell carcinoma, a single-centre retrospective Australian study. Journal of Robotic Surgery. 2024;18(1):226.

14. Faraji F, Kumar A, Voora R, Soliman SI, Cherry D, Courtney PT, et al. Transoral Surgery in HPV‐Positive Oropharyngeal Carcinoma: Oncologic Outcomes in the Veterans Affairs System. The Laryngoscope. 2024;134(1):207–14.

15. Daniels KE, Awad DR, Liu SX, Mocharnuk J, Kubik M, Kim S, et al. Impact of post-operative transoral robotic surgery hemorrhage on adjuvant treatment delays in patients with oropharyngeal squamous cell carcinoma. Oral Oncology. 2024; 159:107031.

16. Lee E, Gorelik D, Crowder HR, Badger C, Schottler J, Li N-W, et al. Swallowing function following neoadjuvant chemotherapy and transoral robotic surgery for oropharyngeal carcinoma: a 2-year follow-up. Otolaryngology–Head and Neck Surgery. 2022;167(2):298–304.

17. Kumar S, Laugharne D, Mortimore S. Revision Transoral Robotic Surgery for Early-Stage HPV Positive Oropharyngeal Squamous Cell Carcinoma, it’s Timing and Margins: Past and Present-a Prospective Single Centre Observational Study. Indian Journal of Otolaryngology and Head & Neck Surgery. 2022;74(Suppl 3):6236–40.

18. Hardman JC, Holsinger FC, Brady GC, Beharry A, Bonifer AT, D’Andréa G, et al. Transoral robotic surgery for recurrent tumours of the upper aerodigestive tract (RECUT): an international cohort study. Journal of the National Cancer Institute. 2022;114(10):1400–9.

19. Ferris RL, Flamand Y, Weinstein GS, Li S, Quon H, Mehra R, et al. Phase II randomized trial of transoral surgery and low-dose intensity modulated radiation therapy in resectable p16+ locally advanced oropharynx cancer: an ECOG-ACRIN cancer research group trial (E3311). Journal of clinical oncology. 2022;40(2):138–49.

20. Nichols DS, Zhao J, Boyce BJ, Amdur R, Mendenhall WM, Danan D, et al. HPV/p16-positive oropharyngeal cancer treated with transoral robotic surgery: The roles of margins, extra-nodal extension and adjuvant treatment. American Journal of Otolaryngology. 2021;42(1):102793.

21. Viros Porcuna D, Granell J, Rama Lopez J, Pollan Guisasola C, TilPerez G. Transoral Robotic Surgery for Squamous Cell Carci-noma of the Oropharynx. Outcomes of the First Multicentric Series in Spain. J Otolaryng Head Neck Surg. 2019; 5:030.

22. Van Abel KM, Quick MH, Graner DE, Lohse CM, Price DL, Price KA, et al. Outcomes following TORS for HPV-positive oropharyngeal carcinoma: PEGs, tracheostomies, and beyond. American Journal of Otolaryngology. 2019;40(5):729–34.

